# Efficacy and safety of early combined therapy with PCSK9 inhibitor and statin in acute ischemic stroke (CAPTAIN): protocol of a multicenter, prospective, open-label, randomized trial

**DOI:** 10.64898/2026.07.27.26358990

**Authors:** Han Qiu, Yi Xie, Siying Xiong, Tingting Qin, Hao Huang, Gang Deng, Chenchen Liu, Yihui Wang, Yi Zhang, Qianqian Kong, Guo Li, Ying Yu, Peixin Li, Thanh Nguyen, Duolao Wang, Shabei Xu, Wei Wang, Xiang Luo

## Abstract

**Background:** Patients with acute ischemic stroke (AIS) attributable to symptomatic intracranial atherosclerotic disease (sICAD) remain at substantial risk of early neurological deterioration (END) despite standard medical treatment.

**Aim:** To determine whether early addition of a proprotein convertase subtilisin/kexin type 9 (PCSK9) inhibitor to statin therapy reduces END in patients with AIS attributable to sICAD within 48 hours of symptom onset.

**Design:** The study of Combined Therapy with PCSK9 Inhibitor and Statin in Acute Ischemic Stroke (CAPTAIN) is a multicenter, prospective, randomized, open-label, blinded-endpoint trial conducted in China, with a planned sample size of 416 patients. Eligible patients will be randomized in a 1:1 ratio to receive either early evolocumab plus standard-dose statin or standard-dose statin therapy.

**Outcomes:** The primary efficacy outcome is the occurrence of END within 3 days after randomization, defined as an increase in the National Institutes of Health Stroke Scale score of at least 2 points from baseline. Secondary efficacy outcomes include neurological improvement, changes in National Institutes of Health Stroke Scale (NIHSS) score, shift in the 90-day modified Rankin Scale score distribution, recurrent stroke, and changes in lipid parameters. The primary safety outcome is the incidence of moderate-to-severe systemic bleeding within 3 days after randomization, as defined by the GUSTO criteria.

**Conclusions:** This study is expected to provide evidence on the efficacy and safety of the early addition of a PCSK9 inhibitor to statin therapy in patients with AIS attributable to sICAD.

**Trial registration number:** NCT06696820

**WHAT IS ALREADY KNOWN ON THIS TOPIC:** Patients with acute ischemic stroke (AIS) attributable to symptomatic intracranial atherosclerotic disease (sICAD) remain at high risk of early neurological deterioration (END) despite standard medical treatment. Previous studies suggested that PCSK9 inhibitors, when added to statin therapy, could rapidly lower LDL-C levels and might reduce END in patients with AIS, but randomized evidence in AIS attributable to sICAD remains limited.

**WHAT THIS STUDY ADDS:** The CAPTAIN trial evaluates the efficacy and safety of the early addition of a PCSK9 inhibitor to statin therapy in patients with AIS attributable to sICAD. This protocol describes the rationale, design, outcomes, and statistical analysis plan of the trial.

**HOW THIS STUDY MIGHT AFFECT RESEARCH, PRACTICE OR POLICY:** CAPTAIN is expected to provide randomized evidence on whether the early addition of a PCSK9 inhibitor to statin therapy reduces END in patients with AIS attributable to sICAD. The findings may inform future research and acute phase lipid-lowering strategies for this high-risk stroke population.

## Introduction

Acute ischemic stroke (AIS) remains a leading cause of death and long-term disability worldwide [1]. Intracranial atherosclerotic disease (ICAD) represents a major cause of ischemic stroke, particularly in Asian populations, where it may account for up to 50% of cases [2,3]. Patients with AIS attributable to symptomatic ICAD (sICAD) are at substantial risk of early neurological deterioration (END) and recurrent ischemic events despite standard medical treatment [3–5]. END is concentrated in the earliest phase of AIS, with a large prospective multicenter cohort showing that 62.5% of END events occurred within 24 hours after admission and that the incidence declined substantially thereafter [6]. Given the close association of END with poor functional recovery and subsequent vascular events [7,8], preventing END represents an important therapeutic target in the acute phase of symptomatic sICAD-related AIS.

Proprotein convertase subtilisin/kexin type 9 (PCSK9) monoclonal antibodies, including evolocumab, produce rapid and intensive reductions in low-density lipoprotein cholesterol (LDL-C) by enhancing hepatic LDL receptor recycling and LDL-C clearance. Beyond lipid lowering, PCSK9 inhibition may also modulate vascular inflammation, endothelial dysfunction, thrombotic activity, and plaque stability [9]. These processes are biologically relevant to sICAD-related AIS, in which culprit-plaque instability, thrombus progression, impaired distal perfusion, and microcirculatory dysfunction may contribute to END [4,10–13]. In the FOURIER trial, evolocumab was added to background statin therapy for long-term secondary prevention in patients with stable atherosclerotic cardiovascular disease, excluding those with myocardial infarction or stroke within the preceding 4 weeks. During a median follow-up of 2.2 years, evolocumab reduced ischemic stroke without a statistically significant increase in hemorrhagic stroke [14]. Similar findings were reported with alirocumab in the ODYSSEY OUTCOMES trial [15]. A prospective multicenter cohort study evaluated early add-on PCSK9 inhibitor therapy in patients with AIS attributable to symptomatic intracranial atherosclerotic stenosis (sICAS) presenting within 7 days of onset. The primary outcome was recurrent stroke within 1 month, which occurred in 2.16% of patients in the PCSK9 inhibitor group and 5.59% of controls (adjusted HR, 0.335; 95% CI, 0.114–0.986) [5]. Despite these findings, adequately powered multicenter randomized controlled trials (RCTs) of early PCSK9 inhibitor therapy in patients with AIS attributable to sICAD are lacking.

To address this evidence gap, we initiated the Combined Therapy with PCSK9 Inhibitor and Statin in Acute Ischemic Stroke (CAPTAIN) trial, a prospective, multicenter, randomized, open-label, blinded-endpoint trial designed to evaluate whether early evolocumab plus statin therapy reduces END in patients with AIS attributable to sICAD within 48 hours after symptom onset.

## Methods

### Design

The CAPTAIN trial is a prospective, multicenter, randomized, open-label, blinded-endpoint (PROBE) trial. The trial was registered at ClinicalTrials.gov (NCT06696820) in November 2024 and has been approved by the Ethics Committee of Tongji Medical College, Huazhong University of Science and Technology (IRB approval number: 2024-S151). Written informed consent must be obtained from all patients or their legal representatives before any study-related procedures are performed. The overall study design is illustrated in **Figure 1**, and the detailed schedule of enrollment, interventions, and assessments is provided in **Supplementary Figure 1**.

**Figure 1.**
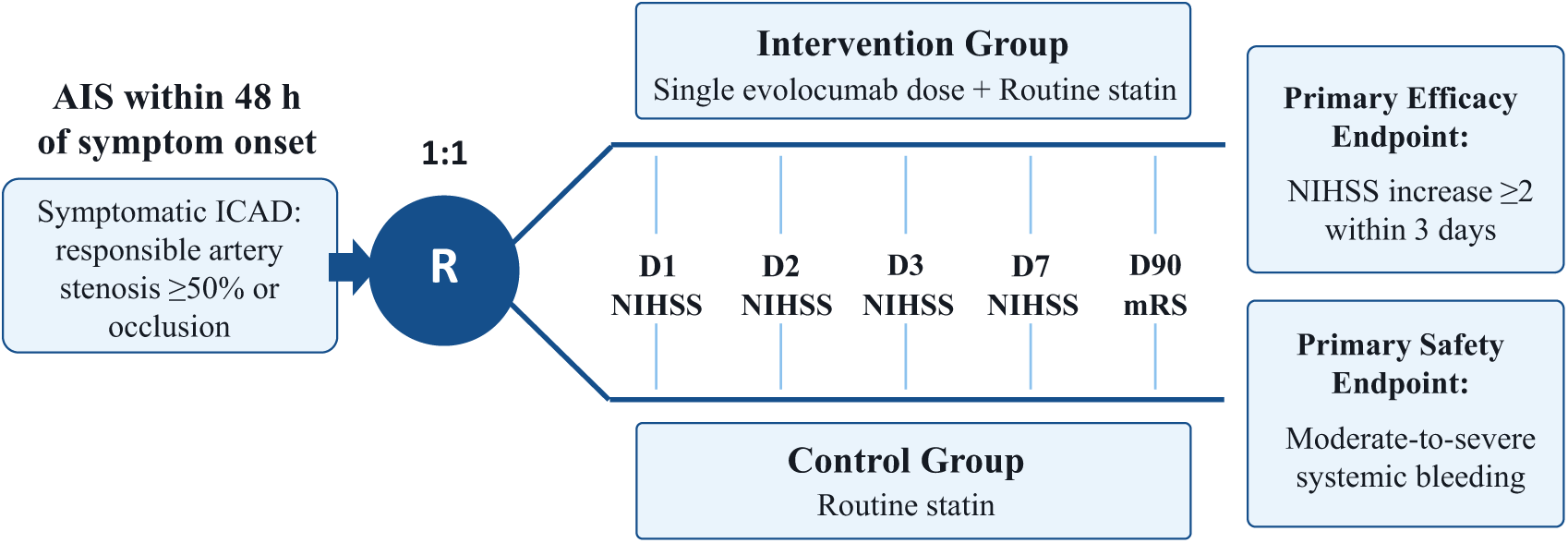
Study design. AIS, acute ischaemic stroke; D1, day 1; D2, day 2; D3, day 3; D7, day 7; D90, day 90; ICAD, intracranial atherosclerotic disease; NIHSS, National Institutes of Health Stroke Scale; mRS, modified Rankin Scale; R, randomisation.

### Study Population

Patients are enrolled from approximately 37 clinical centers across China, beginning in November 2024. The geographic distribution of participating centers is shown in **Supplementary Figure 2**. Trained investigators screen and enroll eligible patients with AIS attributable to sICAD within 48 hours after symptom onset according to standardized procedures. For this trial, sICAD is defined as AIS attributable to 50% to 99% atherosclerotic stenosis or atherosclerotic occlusion of the responsible major intracranial artery, as demonstrated by computed tomography angiography (CTA), magnetic resonance angiography (MRA), or digital subtraction angiography (DSA). Attribution requires an acute infarct in the corresponding vascular territory and the exclusion of cardioembolic and prespecified non-atherosclerotic causes. The inclusion and exclusion criteria are summarized in **Figure 2**. Baseline vascular and brain imaging used to establish sICAD is centrally reviewed by an Imaging Review Committee blinded to treatment allocation to confirm the culprit artery, degree of intracranial stenosis or presence of occlusion, and corresponding infarct territory.

**Figure 2.**
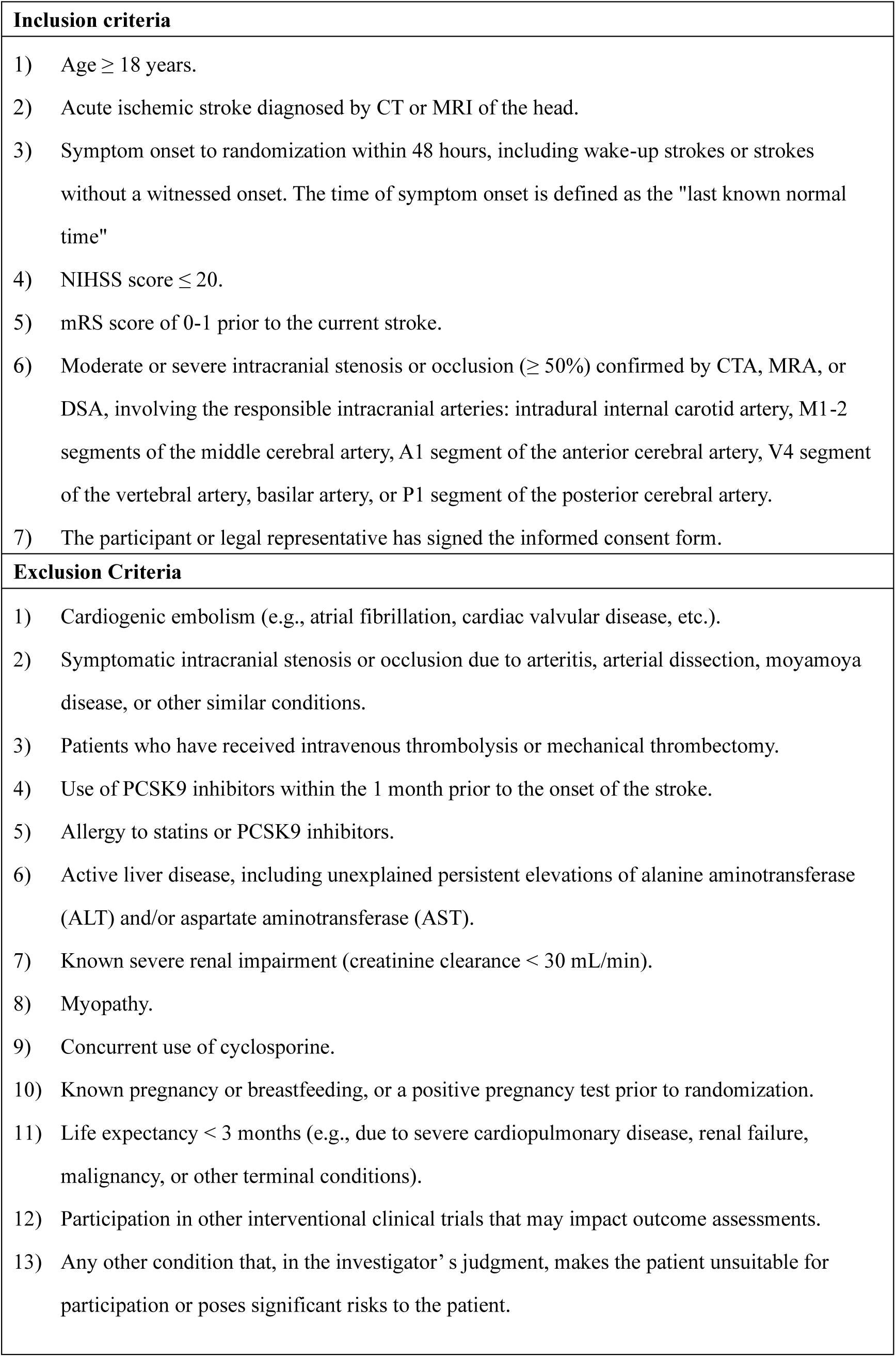
Inclusion and exclusion criteria.

### Randomization

Eligible patients are randomly assigned in a 1:1 ratio to either the PCSK9 inhibitor plus statin group or the statin-alone group through a 24-hour, real-time, centralized web-based system. Randomization is stratified by clinical center and performed using permuted blocks with a block size of 4. Because of the open-label design, treatment allocation is not masked to the treating investigators or participants. However, efficacy outcomes are assessed by qualified outcome assessors who are blinded to treatment allocation.

### Intervention

Patients in the experimental group receive a single subcutaneous injection of evolocumab 420 mg as soon as possible after randomization and within 48 hours after symptom onset, in addition to standard-dose statin therapy with atorvastatin 20 mg or rosuvastatin 10 mg. Patients in the control group receive standard-dose atorvastatin 20 mg or rosuvastatin 10 mg. Other guideline-based treatments for AIS are provided in both groups at the discretion of the treating physicians [16]. Excessive or rapid blood pressure reduction that might compromise cerebral perfusion is avoided during the acute phase. Treatment modifications, concomitant medications, and adverse events are recorded throughout follow-up.

### Efficacy Outcomes

The primary efficacy outcome is the occurrence of END within 3 days after randomization, defined as an increase in the National Institutes of Health Stroke Scale (NIHSS) score of at least 2 points from baseline. Secondary efficacy outcomes include early neurological improvement at 3 days, defined as a decrease in the NIHSS score of at least 4 points from baseline or a decrease to 0 or 1; changes in the NIHSS score from baseline to 1, 3, and 7 days after randomization or discharge; an increase in the NIHSS score of at least 4 points from baseline within 3 days after randomization; the distribution of modified Rankin Scale (mRS) scores at 90 days; the proportions of patients with mRS scores of 0–1, 0–2, and 0–3 at 90 days; recurrent stroke within 30 and 90 days after randomization; and changes in lipid parameters, including LDL-C, high-density lipoprotein cholesterol (HDL-C), total cholesterol, and triglycerides, from baseline to 7 days after randomization or discharge, whichever occurs earlier.

All NIHSS-based efficacy outcomes and the 90-day mRS score are assessed at predefined time points by trained and certified outcome assessors who are blinded to treatment allocation and are not involved in treatment decisions. Prespecified clinical endpoint events are adjudicated by an independent Clinical Event Committee blinded to treatment allocation.

### Safety Outcomes

The primary safety outcome is moderate-to-severe systemic bleeding within 3 days after randomization, defined according to the GUSTO criteria [17]. Secondary safety outcomes include any systemic bleeding within 3 days after randomization, classified according to the GUSTO criteria; any intracranial hemorrhage (ICH) within 3 days after randomization, assessed according to the ECASS morphological criteria [18]; adverse events and serious adverse events within 90 days after randomization; and all-cause mortality within 90 days.

### Sample Size

The sample size was calculated based on the primary efficacy outcome of END within 3 days after randomization. The assumed event rate in the control group is 23.6%, based on the control arm of the TREND trial, an AIS trial in which patients received standard background medical treatment and END within 3 days occurred in 23.6% of patients [19]. The assumed event rate in the experimental group is 12.5%, based on a propensity score-matched cohort study of patients with AIS due to branch atheromatous disease who were treated with a PCSK9 inhibitor in addition to statin therapy [20]. These assumptions correspond to an absolute risk reduction of 11.1 percentage points. Assuming a two-sided alpha level of 0.05, 80% power, and a 1:1 allocation ratio for comparing two independent proportions, 188 participants are required in each group. Allowing for a 10% loss to follow-up, the planned sample size is set at 208 participants per group, for a total of 416 participants.

### Data Management and Quality Control

Data are collected and managed using a centralized, web-based Electronic Data Capture (EDC) system provided by EmpowerStats Co., Ltd., Wuhan, China. Data quality is ensured through a combination of built-in range checks, logic checks, mandatory fields, data queries, audit trails, and regular monitoring. The Data and Safety Monitoring Board (DSMB), consisting of a neurologist, a lipid specialist, and an independent statistician, periodically reviews trial progress, protocol adherence, and safety data, including serious adverse events. Adverse events are assessed according to the Common Terminology Criteria for Adverse Events (CTCAE), version 5.0 [21]. Based on identified safety concerns, the DSMB has the authority to recommend continuation, modification, or termination of the trial to the Steering Committee.

### Statistical Analysis

All statistical analyses will be conducted using R version 4.3.0 and SAS version 9.4, according to a prespecified statistical analysis plan (SAP) that will be finalized before database lock and final outcome analysis. According to the prespecified intention-to-treat (ITT) principle, all randomly assigned patients will be included in the ITT population. This population will be used for all efficacy analyses. The Per-Protocol Set (PPS) will include participants who complete the protocol-specified treatment without major protocol deviations and will be used for sensitivity analyses. The Safety Analysis Set will include participants who receive at least one dose of study treatment and have at least one post-treatment safety assessment.

Baseline demographics and clinical characteristics will be summarized by treatment group. Continuous variables will be described using means and standard deviations or medians and interquartile ranges, as appropriate. Categorical variables will be summarized as counts and percentages.

The primary efficacy outcome will be analyzed in the ITT population using modified Poisson regression with robust error variances, providing risk ratios (RRs) and corresponding 95% confidence intervals (CIs) reported. Supportive analyses will include the corresponding PPS analysis and models adjusted for prespecified baseline covariates. The secondary binary efficacy outcomes will be analyzed using the same modified Poisson regression approach as the primary efficacy outcome. A win ratio approach will be used to evaluate changes in NIHSS score from baseline to day 1, day 3, and day 7 or discharge, whichever occurs earlier, given the discrete and potentially skewed distribution of NIHSS change scores [22]. Changes in lipid parameters from baseline to day 7 or discharge, whichever occurs earlier, will be analyzed using analysis of covariance (ANCOVA), with baseline lipid levels included as covariates. Log transformation or the win ratio approach may be applied if appropriate. The distribution of the 90-day mRS score will be assessed with proportional-odds ordinal logistic regression, under the assumption that the treatment effect is consistent across all mRS cutoffs. The proportional-odds assumption will be assessed using the Brant test. If the proportional-odds assumption is satisfied, common odds ratios and corresponding 95% CIs will be reported. If the assumption is not met, a win ratio analysis may be performed instead. Safety analyses will be conducted in the Safety Analysis Set. The primary safety outcome, moderate-to-severe systemic bleeding, and the secondary safety outcomes, including any systemic bleeding and any ICH, will be analyzed using modified Poisson regression, with RRs and corresponding 95% CIs reported. Time-to-event outcomes, including recurrent stroke and all-cause mortality, will be analyzed using Kaplan-Meier estimates to generate survival curves, log-rank tests for between-group comparisons, and Cox proportional-hazards models to estimate hazards ratios (HRs) with corresponding 95% CIs. Adverse events and serious adverse events will be summarized descriptively by treatment group, presenting the number and proportion of participants experiencing at least one event. All adjusted analyses will include the following prespecified covariates: age, sex, baseline NIHSS score, infarct territory, and baseline LDL-C level.

Prespecified subgroup analyses will assess whether the treatment effect on END within 3 days after randomization differs across clinically relevant baseline variables, including age, sex, time from symptom onset to randomization, baseline NIHSS score, infarct territory, degree of intracranial artery stenosis or occlusion status, baseline LDL-C level, pre-stroke mRS score, histories of diabetes mellitus, hypertension, and pre-stroke use of lipid-lowering therapy. Subgroup effects will be assessed by including treatment-by-subgroup interaction terms in the primary modified Poisson regression model.

Secondary outcomes are considered exploratory, and *P* values will be interpreted descriptively without formal adjustment for multiplicity. All statistical tests will be two-sided, and *P* < 0.05 will be considered statistically significant.

## Discussion

Patients with AIS attributable to sICAD remain at substantial risk of END during the acute phase [2,4]. This early clinical instability underscores the need for therapies beyond conventional antiplatelet treatment and vascular risk factor control [2]. Early addition of a PCSK9 inhibitor is biologically plausible in this setting because of its rapid LDL-C lowering effect and potential effects on plaque stability, vascular inflammation, and thrombotic processes [2,9–12,23–25]. To our knowledge, the CAPTAIN trial is the first and largest multicenter RCT evaluating early evolocumab added to statin therapy for preventing END in patients with AIS attributable to sICAD.

The CAPTAIN trial has several distinctive features compared with previous studies of PCSK9 inhibitors in atherosclerotic cardiovascular disease or acute ischemic stroke. Large cardiovascular outcome trials, including FOURIER and ODYSSEY OUTCOMES, have established the benefit of PCSK9 inhibitors for secondary prevention in patients with established atherosclerotic cardiovascular disease, including a reduction in ischemic stroke risk without a clear increase in hemorrhagic stroke risk [14,15]. However, these trials were not designed to evaluate PCSK9 inhibitor therapy during the acute phase of ischemic stroke or to prevent END. More recently, a six-center open-label RCT in patients with non-cardiogenic AIS suggested that early evolocumab added to atorvastatin reduced END compared with atorvastatin alone [26], with a post hoc exploratory analysis showing a potentially greater effect in atherosclerotic AIS [27]. This finding is biologically plausible because PCSK9 inhibitors may affect lipid-driven plaque progression, vascular inflammation, and thrombotic activity, mechanisms particularly relevant to large-artery atherosclerotic stroke. However, that trial enrolled a broad non-cardiogenic AIS population with heterogeneous stroke mechanism. High-quality randomized evidence remains limited on whether early PCSK9 inhibitor therapy can reduce END in patients with sICAD-related AIS. CAPTAIN was initiated to address this evidence gap, with a focus on imaging-adjudicated sICAD and enrollment across approximately 37 clinical centers with a larger planned sample size.

A key design choice in CAPTAIN is the use of END within 3 days after randomization as the primary efficacy outcome. This outcome is not intended to replace 90-day functional assessment but to capture neurological worsening during the period most closely aligned with the timing of the intervention. Prospective cohort studies have indicated that neurological deterioration occurs predominantly during the early phase after stroke and predicts poor long-term functional outcomes [6–8]. In AIS attributable to sICAD, early worsening may be driven by plaque instability, in situ thrombosis, artery-to-artery embolism, hemodynamic compromise, impaired distal perfusion, or perforator involvement [2,10–13]. These mechanisms support evaluating the early neurological effects of PCSK9 inhibition in this pathophysiologically relevant population. Secondary outcomes, including neurological improvement at 3 days, 90-day mRS, recurrent stroke, and lipid parameters, further assess neurological recovery, functional outcomes, recurrent vascular events, and other biological responses.

Several design features strengthen the CAPTAIN trial. The multicenter randomized design across approximately 37clinical centers and the planned enrollment of 416 participants are expected to improve recruitment efficiency and enhance the representativeness of patients with sICAD-related AIS in routine practice. Although treatment assignment is open label, endpoint assessment is performed by trained assessors blinded to treatment allocation to reduce outcome assessment bias. Neurological and functional outcomes, including NIHSS and mRS assessments, are evaluated using standardized procedures at prespecified time points. The use of an ITT population for the primary efficacy analysis, supportive Per-Protocol analyses, and prospective safety assessment will further support the methodological rigor of the trial while maintaining feasibility in the acute stroke setting. In addition, extended follow-up after randomization is planned to explore longer-term functional status, recurrent stroke, and other vascular events beyond the prespecified core study follow-up.

Several limitations should be acknowledged. First, although END is clinically meaningful and prognostically important, it remains a surrogate rather than a direct measure of long-term disability. Although 90-day functional outcomes will be assessed as secondary outcomes, the trial was not primarily powered to detect differences in long-term functional recovery. Second, the trial excludes patients who received intravenous thrombolysis or endovascular therapy before enrollment; therefore, the findings cannot be generalized to patients treated with acute reperfusion therapy. Third, the trial does not systematically assess postdischarge lipid-lowering therapy or long-term LDL-C goal attainment. Consequently, the durability of the early lipid-lowering effect and its relationship with longer-term clinical outcomes cannot be fully evaluated. Finally, as the study population is recruited entirely from China, where ICAD is relatively common, the generalizability of the findings to other racial and ethnic groups may require further validation.

## Conclusion

CAPTAIN is a multicenter randomized PROBE trial designed to determine whether early evolocumab plus statin therapy can reduce END in patients with AIS attributable to sICAD. By focusing on a high-risk large-artery atherosclerotic stroke population during the early period of neurological instability, CAPTAIN is expected to provide randomized evidence to inform the role of PCSK9 inhibitor-based intensive lipid lowering in this clinical setting.

## Acknowledgements

We are grateful to all the participants in the CAPTAIN program for their contributions to this study.

## Contributors

XL, SBX and WW designed the study; YX, HQ, SYX, HH and TQ drafted the manuscript; DW, TN, GD, CL, YW, YZ, QK, GL, YY and PL provided critical comments/revisions of the manuscript. XL is the guarantor.

## Patient consent for publication

Not applicable.

## Ethics approval

This study involves human participants and was approved by the Ethics Committee of Tongji Medical College, Huazhong University of Science and Technology (IRB approval number: 2024-S151). Written informed consent must be obtained from all patients or their legal representatives.

## Data availability statement

Data are available upon reasonable request.

