## Supplementary Figure 1and2 for "Efficacy and safety of early combined therapy with PCSK9 inhibitor and statin in acute ischemic stroke (CAPTAIN): protocol of a multicenter, prospective, open-label, randomized trial"

**Supplementary Figure 1 Schedule of enrollment, interventions, and assessments.**

| | | **TRIAL PERIOD** | | | | | |
|---|---|---|---|---|---|---|---|
| | **Enrollment** | **Post-randomization** | | | | | **Close-out** |
| TIMEPOINT | 0 | 24±12 hours | 48±12 hours | 72±12 hours | 7±1days/ discharge | 30±3 days | 90±7 days |
| **ENROLLMENT:** | | | | | | | |
| Eligibility screen | X | | | | | | |
| Informed consent | X | | | | | | |
| Randomization | X | | | | | | |
| **INTERVENTION:** | | | | | | | |
| PCSK9 inhibitor plus statin | X | | | | | | |
| Statin alone | X | | | | | | |
| **ASSESSMENTS:** | | | | | | | |
| Demographics | X | | | | | | |
| Present illness | X | | | | | | |
| Medical history | X | | | | | | |
| mRS | X[#] | | | | | | X |
| NIHSS | X | X | X | X | X | | |
| CT/MRI | X | | | X* | | | |
| CTA/MRA/DSA | X | | | | | | |
| Lipid profile | X | | | | X | | |
| Other laboratory tests | X | | | | X | | |
| ECG | X | | | | | | |
| AE/SAE | | X | X | X | X | X | X |
| Telephone follow-up | | | | | | X | X |

[#] Pre-stroke mRS.

* Brain CT or MRI may be performed if available.

ECG, electrocardiogram; mRS, modified Rankin Scale; NIHSS, National Institutes of Health Stroke Scale; CT, computed tomography; MRI, magnetic resonance imaging; CTA, computed tomography angiography; MRA, magnetic resonance angiography; DSA, digital subtraction angiography; AE, adverse event; SAE, serious adverse event.

**Supplementary Figure 2 Distribution map of study sites.**

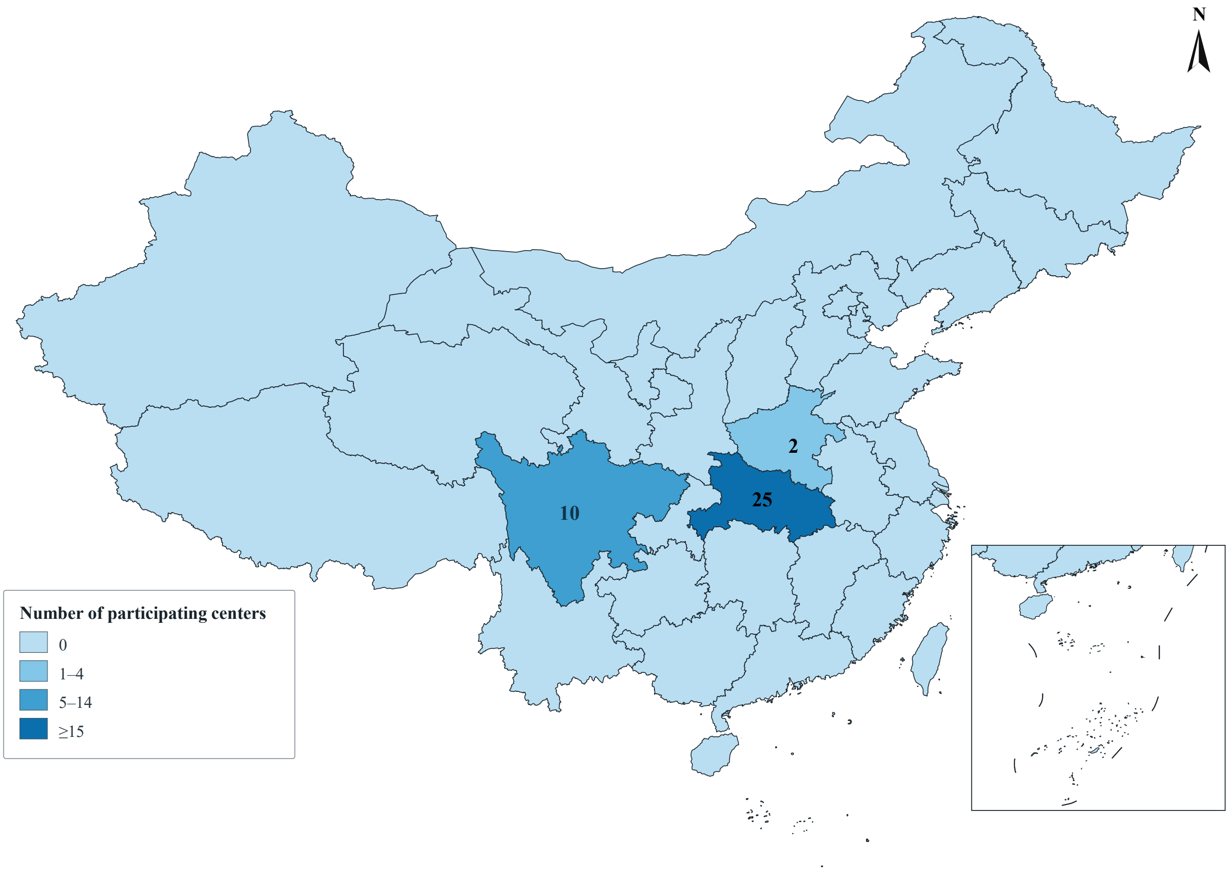


The areas illuminated in blue represent the sites, and the numbers indicate the number of centers.
